# Clinical Characteristics and Factors Associated with Long COVID in Zambia, August 2020 to January 2023: A Mixed Methods Design

**DOI:** 10.1101/2024.01.17.24301423

**Authors:** Warren Malambo, Duncan Chanda, Lily Besa, Daniella Engamba, Linos Mwiinga, Mundia Mwitumwa, Peter Matibula, Neil Naik, Suilanji Sivile, Simon Agolory, Andrew Auld, Lloyd Mulenga, Jonas Z. Hines, Sombo Fwoloshi

## Abstract

**Introduction:** A number of seroprevalence studies in Zambia document the extent of spread of SARS-CoV-2, yet few have examined signs, symptoms and conditions that continue or develop after acute COVID-19 infection (long COVID). This is an important gap given the estimated prevalence of long COVID in other countries. We sought to examine characteristics of post-acute COVID-19 (PAC-19) clinics patients in Zambia and assess factors associated with long COVID at first visit to a PAC-19 clinic and longitudinally among a cohort of patients.

**Methods:** Long COVID was defined, initially in the Zambia PAC-19 clinical guidelines, as new, relapsing, or persistent symptoms lasting >4 weeks after an initial SARS-CoV-2 infection. Severe COVID-19 was defined as COVID-19 episode that required supplemental oxygen therapy, intensive care unit stay or treatment with steroids/remdesivir. We performed a cross-sectional and longitudinal analysis of PAC-19 clinic patients from August 2020 to January 2023 using logistic and mixed effects regression models and considered statistical significance at p<0.05.

**Results:** In total, 1,359 patients attended PAC-19 clinics of whom 548 (40.3%) with ≥2 visits were included in the longitudinal analysis. Patients’ median age was 53 (interquartile range [IQR]: 41-63) years, 919 (67.6%) were hospitalized for acute COVID-19, and of whom 686 (74.6%) had severe COVID-19. Patients with hospital length of stay ≥15 days (adjusted odds ratio [aOR]: 5.37; 95% confidence interval [95% CI]: 2.99-10.0), severe illness (aOR: 3.22; 95% CI: 1.68-6.73), and comorbidities (aOR:1.50; 95% CI: 1.02-2.21) had significantly higher likelihood of long COVID. Longitudinally, long COVID prevalence significantly (p<0.001) declined from 75.4% at the first PAC-19 visit to 26.0% by the fifth visit. The median follow-up time was 7 (IQR: 4-12) weeks.

**Conclusion:** Long COVID symptoms were common among patient presenting for care in PAC-19 clinics in Zambia, but most recovered within ∼2 months. Despite potentially substantial morbidity due to long COVID, few patients overall with COVID-19 attended a PAC-19 clinic. Scaling up PAC-19 services and integrating into routine clinical care could improve access by patients.

## Introduction

Since the identification of the first SARS-CoV-2 infections in December 2019, the Coronavirus disease 2019 (COVID-19) has become a major public health problem. Its morbidity and mortality dramatically increased across countries and regions leading to its classification as a pandemic [1]. As of February 2024, the World Health Organization (WHO) had reported over 774 million cumulative COVID-19 cases and over 7 million cumulative COVID-19 deaths globally [2]. In the Africa region alone, over 9.5 million cumulative COVID-19 cases and over 175 thousand COVID-19 deaths were reported for the same time period.

People infected with SARS-CoV-2 commonly develop symptoms within 4-5 days following exposure although others may be asymptomatic [3]. Acute COVID-19 (within 4 weeks from the initial SARS-CoV-2 infection) presents with as a multi-system cluster of symptoms, i.e., general, cardiovascular, pulmonary, gastrointestinal, neurologic, musculoskeletal, and ear, nose, and throat. Recovery from COVID-19 for most patients occurs within 7-10 days after symptoms onset but could take weeks to months in some patients [4–7]. Previous epidemics such the 2003 severe acute respiratory syndrome (SARS) and the 2012 Saudi Arabia Middle East respiratory syndrome coronavirus (MERS-CoV) had patients who presented with persistent symptoms post-acute infection [8–11]. Their symptoms included fatigue, decreased quality of life, shortness of breath and behavior health problems that impacted on their health-related quality of life.

Symptoms and conditions that continue or develop after acute COVID-19 infection are variably named (post-acute COVID-19, post-acute sequelae of SARS-CoV-2 infection, post-COVID conditions, long-haul COVID, post-COVID conditions, long-term effects of COVID and chronic COVID) but are also referred to as long COVID [6, 12]. Commonly reported long COVID symptoms include fatigue, fever, cough, dizziness, brain fog, and myalgia although symptoms are varied and potentially have overlapping etiologies [13–15]. Long COVID is thus a syndrome characterized by persistent, new, relapsing or delayed SAR-CoV-2 symptoms ≥12 weeks beyond onset of the acute episode [12, 16–18], and can have a protracted path to recovery that impacts on quality of life, earnings and health care costs [19–21].

As much as 77 million people around the world could be estimated to have long COVID, based on a conservative estimated prevalence of 10% of reported number of infected people [2, 22]; the actual number may likely be much higher given undocumented cases due to limited testing capacity. Among acute-COVID-19 outpatient cases, the prevalence of long COVID is estimated at 10-30%; 50-70% of hospitalized cases and 10-12% of vaccinated cases [22–27]. For example, a study of the burden, causation, and particularities of long COVID in 7 African countries estimated a pooled long COVID prevalence of 41% [28]. Long COVID is associated with increasing age, comorbidities, hospitalization for acute COVID-19, severe COVID-19, and vaccination status [29–31]. Vaccination against SARS-CoV-2 is, however, associated with reduced likelihood of severe COVID-19 and long COVID.

There’s growing evidence on long COVID from countries in Africa [28, 32–34]. Disease severity and admission to the intensive care unit during acute COVID-19, for example, were found to be associated with long COVID in Nigeria and South Africa [35, 36]. Scoping studies on long COVID in Africa found comorbidities and age >40 years to be associated factors [33, 37]. In Zambia, 17% of persons with acute COVID-19 in July 2020 were found to be experiencing symptoms ∼2 months later [38]. Zambia’s reported COVID-19 cases are, however, likely an underestimate given limited testing capacity, asymptomatic infections, and mild clinically ill cases who may not have come to the attention of the health system [39–41].

While a number of seroprevalence studies in Zambia document the extent of spread of SARS-CoV-2, few have examined longer term outcomes such as long COVID [36, 39, 41–45]. This is an important gap given the estimated prevalence of long COVID and extent of SARS-CoV-2 transmission in other countries in Africa. In this study, we examined the characteristics of patients presenting for care in specialized post-acute COVID-19 (PAC-19) clinics in Zambia and assessed factors associated with long COVID at first visit to a PAC-19 clinic and longitudinally among a cohort of patients with ≥2 PAC-19 clinic visits.

## Methods

### Study setting

Beginning August 2020, the Zambia Ministry of Health set up 13 specialized PAC-19 clinics to care for people following SARS-CoV-2 infection, initially at the two national referral hospitals in Lusaka city, the capital of Zambia, and subsequently at 11 other major hospitals across all 10 provinces of Zambia.

### Study participants

Per national PAC-19 clinical guidelines, patients diagnosed with SARS-CoV-2 infection presented in PAC-19 clinics for follow-up care after discharge from hospital admission or outpatient episodic [46]. At the first PAC-19 clinic visit, patients’ demographics, medical history, comorbidities, and current symptoms were recorded on a standardized paper form. Physical examinations and laboratory investigations were conducted based on patient’s medical history as required. Patients’ ability to perform activities, as part of examination of their functional and mental health status, were also evaluated. Further clinical appointments of up to 5 review visits were at the clinicians’ discretion. Symptoms were assessed by clinicians at each PAC-19 clinic visit on review of systems: general, cardiovascular, pulmonary, urinary, neurologic, musculoskeletal, ear, nose, and throat (ENT), mental health concerns, depression, anxiety, and dermatologic. Pre-existing comorbidities (hypertension, diabetes, cardiovascular disease, cancer, chronic lung disease, kidney disease, or liver disease, immunosuppression, obesity, HIV and TB) were documented. Patients requiring further specialist care were referred to various units including physiotherapy, cardiology, endocrinology, nephrology, psychiatry, and pulmonology. Anonymized data from patients standardized clinical paper forms were routinely abstracted into an electronic database (REDCap v11.0.3); which was accessed on 5^th^ January 2023 and analyzed for this study.

### Study design

We implemented a mixed methods design to assess for factors associated with long COVID at first visit to a PAC-19 clinic and longitudinally across review visits. First, we performed a cross-sectional analysis of all PAC-19 clinic patients with data entered in the REDCap electronic databases from August 5, 2020, through to January 26, 2023. We then did a longitudinal sub-analysis of patients with ≥2 PAC-19 clinic review visits.

The longitudinal sub-analysis data structure included repeated observations of long COVID symptoms and patient-level characteristics. Demographic and clinical characteristics of patients included time-invariant covariates sex, age (patients repeatedly observed for <1 year), presence of newly diagnosed medical conditions, presence of pre-existing comorbidities, and acute COVID-19 episode details. Time-variant covariates included COVID-19 vaccination status and referral to specialist services.

### Study variables and definitions

Long COVID was defined, initially in the Zambia post-acute COVID-19 guidelines, as new, relapsing or persistent COVID-19 symptoms that lasted greater than 4 weeks after the initial SARS-CoV-2 infection [46, 47]. Severe COVID-19 was defined as COVID-19 episode that required supplemental oxygen therapy, intensive care unit (ICU) stay or treatment with steroids or remdesivir. Vaccination status categorization was based on vaccination records, when available, or patients’ self-reported status. SARS CoV-2 variants classification was based on the dominant variant at the time of SARS-CoV-2 diagnosis as captured by the Zambia’s genomic surveillance system and submitted to the Global Initiative on Sharing All Influenza Data (GISAID) rather than sequenced specimens from patients [48]. Diabetes, hypertension, or deep vein thrombosis/pulmonary embolism detected at the time of SARS-CoV-2 diagnosis were categorized as newly diagnosed medical conditions.

### Study size

For the study sample size, we assumed prevalence of long COVID at 30% and a precision corresponding to the effect size of 5%. The prevalence or Cochrane formula (described elsewhere) was used to calculate the minimum sample size of 323 [49]. Study participants were included in the study if they were COVID-19 patients presenting in PAC-19 clinics for follow-up care after discharge from hospital or outpatient episodic care and had their information abstracted to the clinics’ REDCap electronic database. A full enumeration of all PAC-19 clinic attendees was however considered for higher accuracy and precision. For the longitudinal sub-analysis, patients were excluded from the analysis if they attended a PAC-19 clinic only once.

### Statistical Analysis

At descriptive statistics we reported frequencies with proportions for categorical variables and medians with the interquartile range (IQR) for non-normally distributed numeric variables assessed using the Shapiro-Wilk test. The Pearson Chi-square and Kruskal Wallis tests for proportions and Wilcoxon rank sum test for medians were used to estimate the direction and magnitude of association. An unadjusted and adjusted logistic regression model was fitted to assess for factors associated with having long COVID at first visit to a PAC-19 clinic.

The main cross-sectional analysis was for patients hospitalized during acute COVID-19 (inpatients) to maintain key variables in the analysis. At multivariable analysis, we adjusted for age, presence of newly diagnosed medical conditions, presence of pre-existing comorbidities, and COVID-19 episode details (i.e., hospital length of stay, and presence of severe COVID-19). Covariate inclusion criteria at multivariable analysis were based on theoretical relevance to the study and statistical significance (p-value≤0.2). We also conducted an additional cross-sectional analysis that included both inpatients and outpatients during acute infection to assess for association of hospitalization during acute COVID-19 with long COVID.

For the longitudinal sub-analysis, we a fitted mixed effects model to account for the heterogeneity in long COVID i.e., the differences between patients and within patients across repeated observations or PAC-19 clinic visits. An unadjusted and adjusted mixed effects model was separately fitted for acute COVID-19 inpatients only and additionally for both inpatients and outpatients. The patient was the random effects term for the model. Since there was dependence in the data, we also reported the conditional intraclass correlation (ICC) which quantified the variance in long COVID heterogeneity that was attributable to between patients and within patients across PAC-19 visits.

Only covariates with <10% missingness were included at multivariable analyses. To assess for bias, we checked for the proportion of listwise deletion when covariates of interest with >10% missingness were included at multivariable analysis. Model selection was based on goodness-of-fit statistics (i.e., adjusted pseudo R^2^, Akaike information criteria, Bayesian information criteria and log-likelihood value). Analysis was done in R software version 4.3.2 (R Foundation for Statistical Computing) and statistical significance was considered at p<0.05.

### Ethical considerations

This study was preconceived during the development of clinical and operational guidelines for the management of PAC-19 patients in Zambia. The study obtained ethical clearance (waiver for informed consent) from the University of Zambia Biomedical Research Ethics Committee (Ref No. 2711-2022), approval from the Zambia National Health Research Authority (Ref No: NHRA0002/26/05/2022) and was determined to be non-research according to the U.S Centers for Disease Control and Prevention (CDC) policy and applicable federal law (See e.g., 45 C.F.R. part 46.102(l)(2), 21 C.F.R. part 56; 42 U.S.C. §241(d); 5 U.S.C. 552a; 44 U.S.C.3501 et seq.). CDC investigators did not interact with patients or have access to personally identifiable information but participated in protocol development and analysis of anonymized data.

## Results

In total, 1,359 patients that attended PAC-19 clinics from August 2020 to January 2023 were in the cross sectional analysis (Fig 1). Of these patients, 548 (40.3%) patients that had ≥2 PAC-19 clinic visits were included in the longitudinal sub-analysis. Most patients (919/1,087, 84.5%) were hospitalized during acute COVID-19 (Table 1). Overall, patients’ median age was 53 (IQR: 41-63) years, and 693 (52.9%) were female. Most patients (58.0%) were from Lusaka, where the first two PAC-19 clinics were initially set up. Nearly half of the patients (46.9%) were diagnosed with COVID-19 when delta was the dominant variant (Fig. 2). The median time since COVID-19 diagnosis when patients presented for care in PAC-19 clinics was 4 (IQR:2-6) weeks.

**Fig 1:**
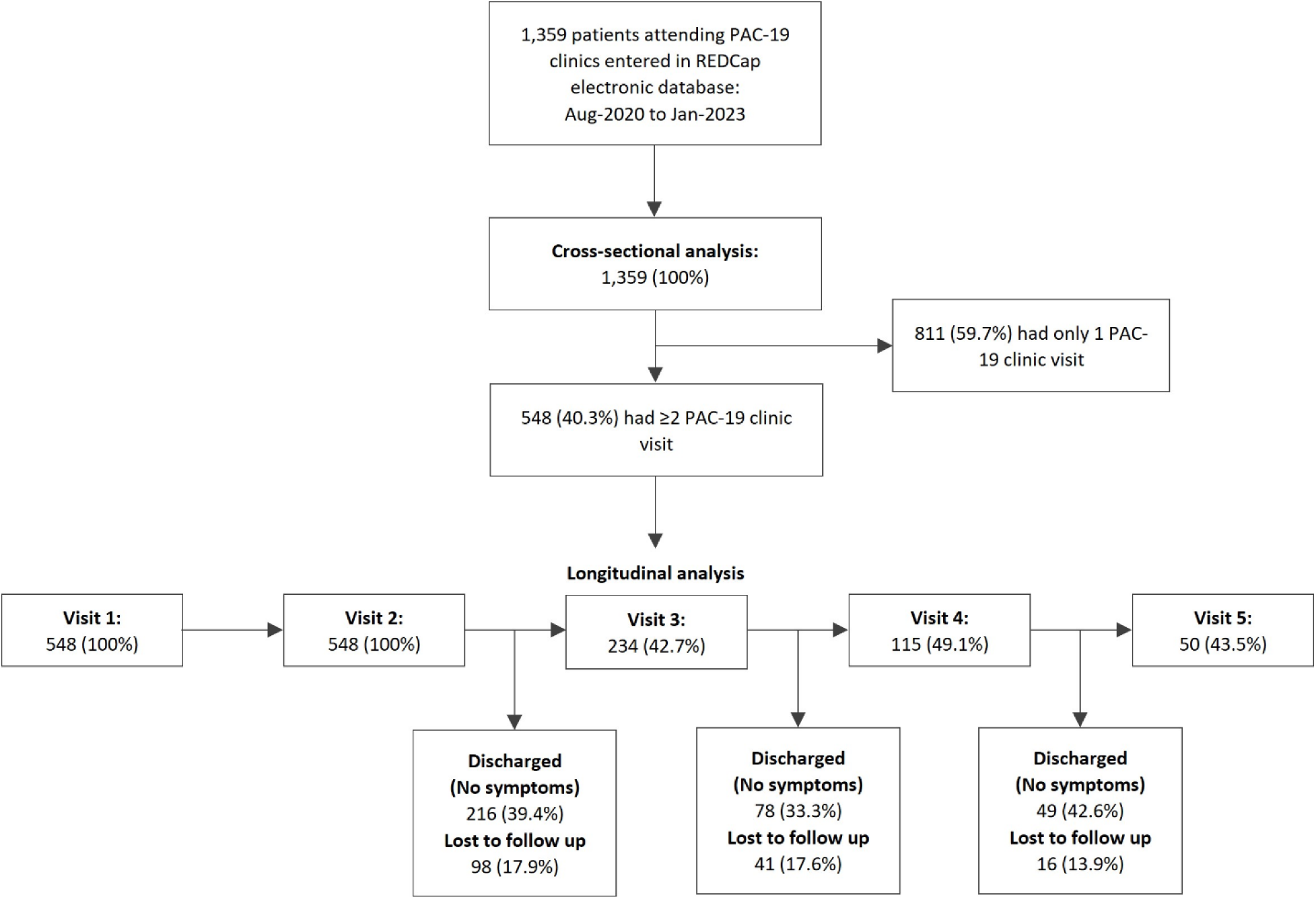
Analysis flow diagram of PAC-19 clinic patients in Zambia, August 2020 – January 2023

**Fig 2:**
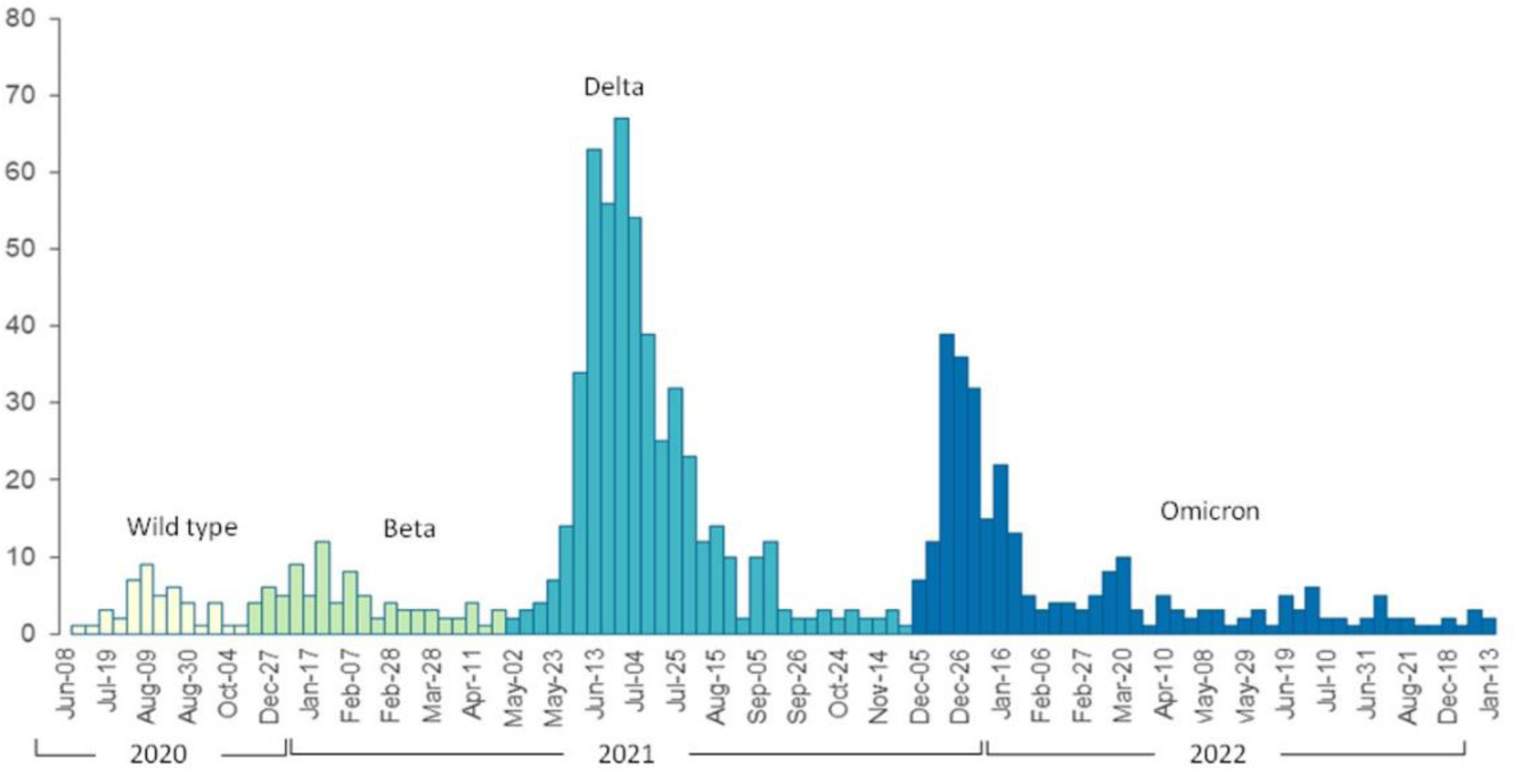
PAC-19 clinic patients’ attendance by date of diagnosis, August 2020 – January 2023

**Table 1:**
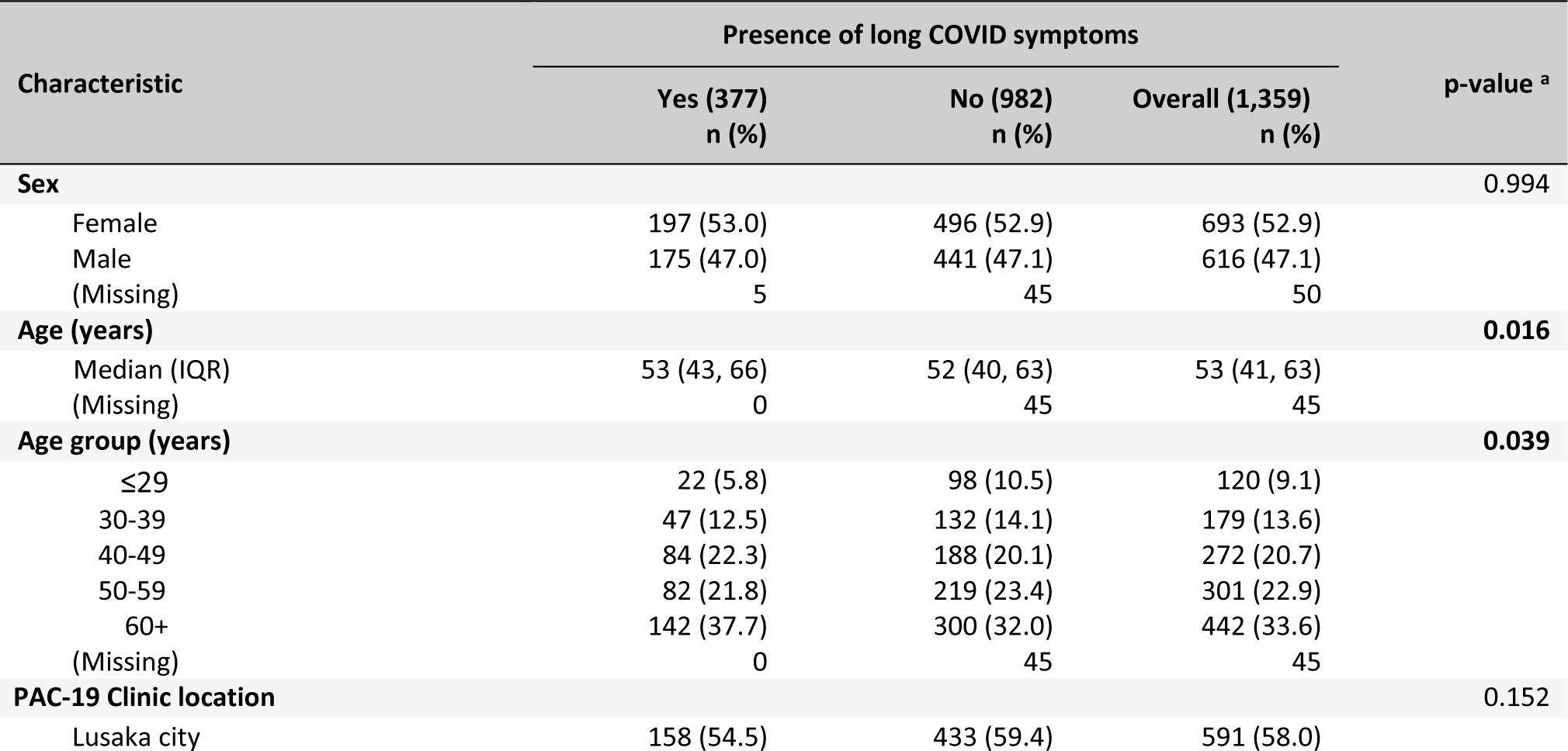
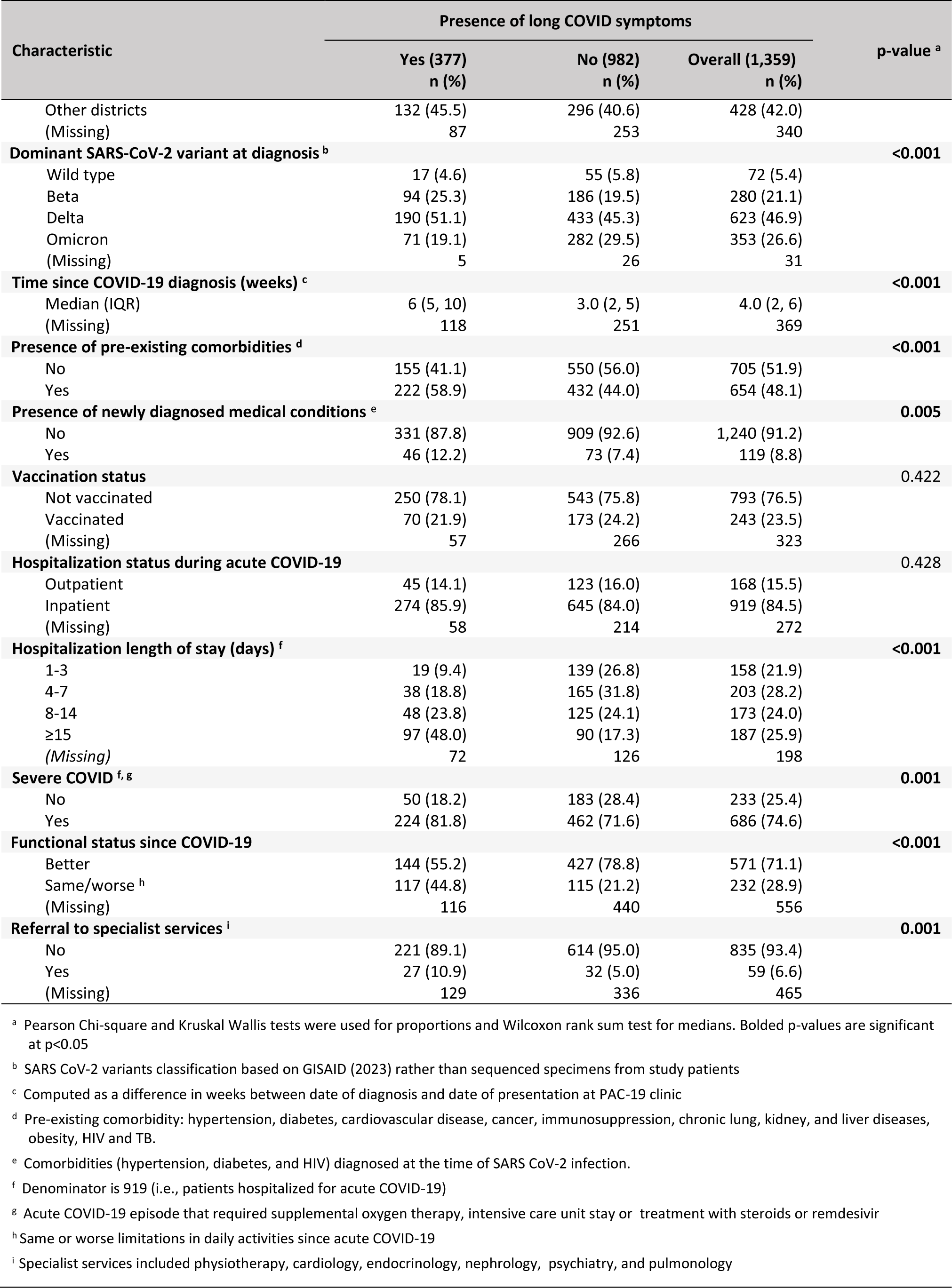
Demographic and Clinical Characteristics of Patients who Presented for Care in PAC-19 Clinics in Zambia, Aug. 2020‒Jan. 2023.

At baseline, 654 (48.1%) patients overall, reported having had pre-existing comorbidity. Among these patients, hypertension (n=465, 71.1%), diabetes (n=172, 26.3%), HIV (n=165, 25.2%) and cardiovascular disease (n=73, 11.2%) were commonly (frequency of ≥10%) reported (S1 Table). One hundred nineteen (8.8%) patients had been diagnosed with new medical conditions at the time of SARS-CoV-2 diagnosis. Of these patients, hypertension and diabetes were detected at SARS-CoV-2 diagnosis in 68 (53.8%) and 64 (53.8%) patients, respectively.

Among the hospitalized acute COVID-19 patients, the median length of stay was 7 days (IQR:4-15 days). Of these inpatients, 686 (74.6%) were categorized as having had severe COVID-19. The medium time from the date of COVID-19 diagnosis to when patients presented for care in a PAC-19 clinic was 4 weeks (IQR: 2-6 weeks). At first visit to a PAC-19 clinic, 274 (29.8%) hospitalized patients had long COVID. Among these long COVID patients, commonly reported symptoms included cough (38.7%), fatigue (38.5%), shortness of breath (26.5%), chest pain (20.9%), headache (14.8%), muscles aches/pain (14.1%), palpitations (12.5%), joint aches/pain (11.9%), and forgetfulness or brain fog (8.0%). Among the outpatients, 103 (23.4%) had long COVID, bringing the overall number of long COVID patients to 377 (27.7%).

Of all patients, 232 (28.9%) patients had worse or same (limitations in daily activities) functional status since acute COVID-19. The functional status patients commonly reported difficulty in undertaking was walking long distances greater than 1km (22.6%), day-to-day work/school (cognitive) activities (19.4%), standing for ≥30 minutes (19.4%), taking care of household tasks (17.8%), and self-care (activities like bathing and dressing) – 15.1%. Fifty-nine (6.6%) patients were referred to specialist care, including cardiology (35.6%), endocrinology (33.9%), psychiatry (25.4%), pulmonology (18.6%), physiotherapy (11.0%), and nephrology (1.7%).

Overall, 243 (23.5%) patients had been vaccinated before being diagnosed with SARS-CoV-2, representing 28.5% of patients seen at PAC-19 clinics from April 2021 when COVID-19 vaccines first became available in Zambia. Of these vaccinated patients, 102 (42%%) received Johnson and Johnson’s Janssen (Ad26.COV2.S), 97 (39.9%%) received AstraZeneca (AZD1222), 41 (16.9%%) did not know the vaccine type they received, and 3 (1.2%%) received Pfizer-BioNTech (BNT162b2). Fifty-seven (23.4%) had received a full series of vaccines >14 days prior to their date of SARS-CoV-2 diagnosis, 31 (12.7%) were partially vaccinated (i.e., 2nd dose not yet received), 10 (4.1%) were diagnosed with SARS-CoV-2 within 14 days of being vaccinations while 148 (60.9%) had missing vaccination dates to be classified.

Patients with pre-existing comorbidities (adjusted odds ratio [aOR]: 1.50; 95% confidence interval [CI]: 1.02-2.21), and with hospital length of stay of 8-14 and ≥15 days were associated with significantly higher odds of long COVID (aOR: 1.98; CI: 95% 1.08-3.74, and aOR: 5.37; 95% CI: 2.99-10.0, respectively) - Table 2. Patients who had severe COVID-19 had 3.23-fold (95% CI: 1.68-6.75) higher odds of presenting with long COVID. Independently, vaccinated patients had non-significant reduced likelihood of long COVID (OR: 0.94; 95% CI: 0.64-1.36) while those who were referred to specialist services had 1.88-fold (95% CI: 1.00-3.48; p-value=0.045) increased likelihood of long COVID.

**Table 2:** Factors associated with long COVID at first visit to PAC-19 clinic among patients hospitalized for acute COVID-19 in Zambia, August 2020 – January 2023 (N=919)

| Variable | Unadjusted OR<br>(95% CI) | p-value <sup>a</sup> | Adjusted OR<br>(95% CI) | p-value <sup>a</sup> |
| --- | --- | --- | --- | --- |
| **Sex** |  |  |  |  |
| Female | Referent | - | Referent | - |
| Male | 0.98 (0.74, 1.31) | 0.912 | 0.92 (0.65-1.32) | 0.667 |
| **Age group (years)** |  |  |  |  |
| ≤29 | Referent | - | Referent | - |
| 30-39 | 1.73 (0.88, 3.51) | 0.116 | 0.87 (0.38, 2.01) | 0.741 |
| 40-49 | 2.01 (1.09, 3.87) | **0.029** | 0.82 (0.39, 1.76) | 0.596 |
| 50-59 | 1.69 (0.93, 3.24) | 0.096 | 0.70 (0.33, 1.51) | 0.350 |
| 60+ | 1.86 (1.05, 3.47) | **0.041** | 0.61 (0.30, 1.28) | 0.181 |
| **Presence of pre-existing comorbidities <sup>b</sup>** |  |  |  |  |
| No | *Referent* |  | *Referent* | - |
| Yes | 1.50 (1.12, 2.01) | **0.006** | 1.50 (1.02, 2.21) | **0.041** |
| **Presence of newly diagnosed medical conditions <sup>c</sup>** |  |  |  |  |
| No | *Referent* | - | *Referent* | - |
| Yes | 1.48 (0.94, 2.30) | 0.084 | 1.34 (0.78, 2.28) | 0.284 |
| **Hospital length of stay (days)** |  |  |  |  |
| 1-3 | *Referent* | - | *Referent* | - |
| 4-7 | 1.68 (0.94, 3.11) | 0.086 | 1.26 (0.68, 2.39) | 0.478 |
| 8-14 | 2.81 (1.59, 5.14) | **<0.001** | 1.98 (1.08, 3.74) | **0.031** |
| ≥15 | 7.88 (4.60, 14.1) | **<0.001** | 5.37 (2.99, 10.0) | **<0.001** |
| **Severe COVID-19 <sup>d</sup>** |  |  |  |  |
| No | *Referent* |  | *Referent* | - |
| Yes | 1.77 (1.26, 2.54) | **0.001** | 3.22 (1.68, 6.73) | **<0.001** |
| **Vaccination status <sup>e</sup>** |  |  |  |  |
| Not vaccinated | *Referent* |  | - | - |
| Vaccinated | 0.94 (0.64, 1.36) | 0.737 | - | - |
| **Referral to specialist services <sup>e</sup>** |  |  |  |  |
| No | *Referent* |  | - | - |
| Yes | 1.88 (1.00, 3.48) | **0.045** | - | - |

Abbreviation: OR - odds ratio; CI – confidence interval
<sup>a</sup> Bolded p-values are significant at p<0.05
<sup>b</sup> Pre-existing comorbidity: hypertension, diabetes, cardiovascular disease, cancer, immunosuppression, chronic lung, kidney, and liver diseases, obesity, HIV and TB.
<sup>c</sup> Comorbidities (hypertension, diabetes, and HIV) diagnosed at the time of SARS CoV-2 infection.
<sup>d</sup> Acute COVID-19 episode that required supplemental oxygen therapy, intensive care unit stay or treatment with steroids/ remdesivir
<sup>e</sup> Vaccination status and referral to specialist services were not adjusted for at multivariable analysis due to >10% missingness which resulted in 50.5% listwise deletion.

In the additional cross-sectional analysis that included both inpatients and outpatients during acute COVID-19 (S2 Table), inpatients were associated with non-significant increased odds of long COVID (aOR: 1.05; 95% CI: 0.72-1.55). Patients with the presence of comorbidities were, however, associated with significantly higher odds of long COVID (aOR: 1.55; 95% CI: 1.16-2.08) while the presence of newly diagnosed medical conditions had a non-significant increase likelihood of long COVID. Patients aged 40-49 years were significantly associated with 1.79-fold increased likelihood of long COVID (95% CI: 1.03-3.22). Independently, patients referred to specialist services were 2.34 times (95% CI: 1.37-4.00) more likely to present with long COVID. Males compared to females and having been vaccinated against COVID-19 were associated with non-significant reduced odds of long COVID.

In the longitudinal analysis (S1 Fig), the overall median time patients presented at a PAC-19 clinic was 7 (IQR: 4-12) weeks. Longitudinally (Fig 3), the most frequently (≥5% at any PAC-19 clinic visit) reported long COVID symptoms were cough (16.9%), fatigue (16.4%), shortness of breath (10.6%), chest pain (8.4%), headache (8.3%), palpitations (6.6%) and muscle aches/pains (5.2%), and forgetfulness (3.0%). The prevalence of long COVID significantly declined (p<0.001) from 75.4% at the first PAC-19 clinic visit to 53.5% at the second visit, 59.8% at the third visit, 49.6% at the fourth visit, and 26.0% at the final visit (S3 Table). Overall, the proportion of vaccinated patients attending PAC-19 clinics increased from 20% at the first visit to 58% at the fifth visit; an increase that will statistically significant (p<0.001). Similarly, comparatively significantly (p=0.002) larger proportion of patients with comorbidities presented in subsequent PAC-19 clinic visits than at the first visit. The proportion of patients with newly diagnosed medical conditions longitudinally declined; this was, however, non-significant (p=0.953).

**Fig 3:**
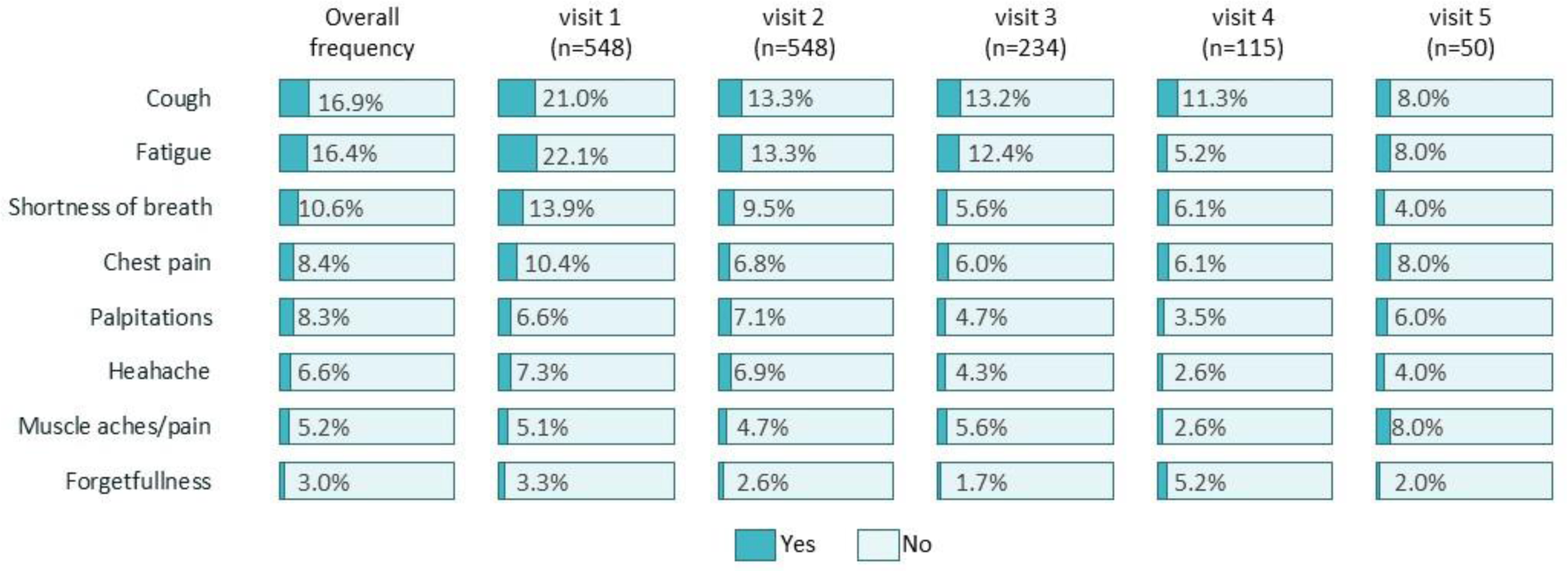
Longitudinal frequency of most common long COVID symptoms by PAC-19 clinic visit, Aug. 2020 - Jan. 2023

We found similar associations with long COVID in the longitudinal analysis (Table 3) as with the cross-sectional analysis (Table 2). Across subsequent clinical visits, the odds of long COVID declined overall. Patients with hospital length of stay of ≥15 days and those with severe COVID-19 had significantly increased likelihood of presenting with long COVID (aOR: 4.30; 95% CI: 1.54-12.0, and aOR: 1.89; 95% CI: 1.02-3.49, respectively). Longitudinally, vaccinated patients had a non-significant reduced odds of long COVID (aOR: 0.78; 95% CI: 0.45-1.35). Similarly, sex, age, presence of comorbidities and newly diagnosed medical conditions were associated with non-significant likelihood of long COVID. For the longitudinal model, the conditional ICC was estimated to be 0.562; suggesting that 56.2% of the longitudinal variance in long COVID was attributable to between patient differences.

**Table 3:**
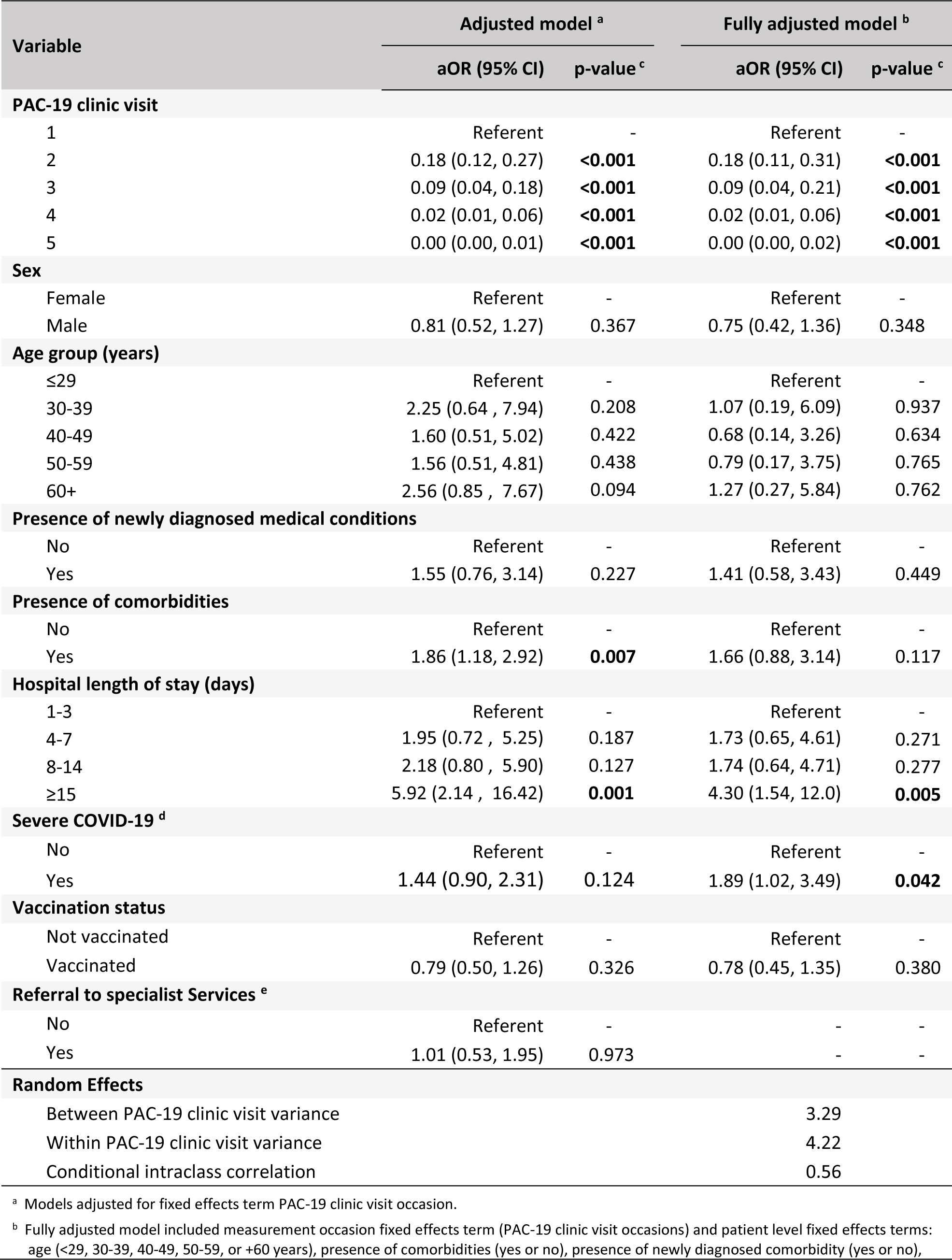
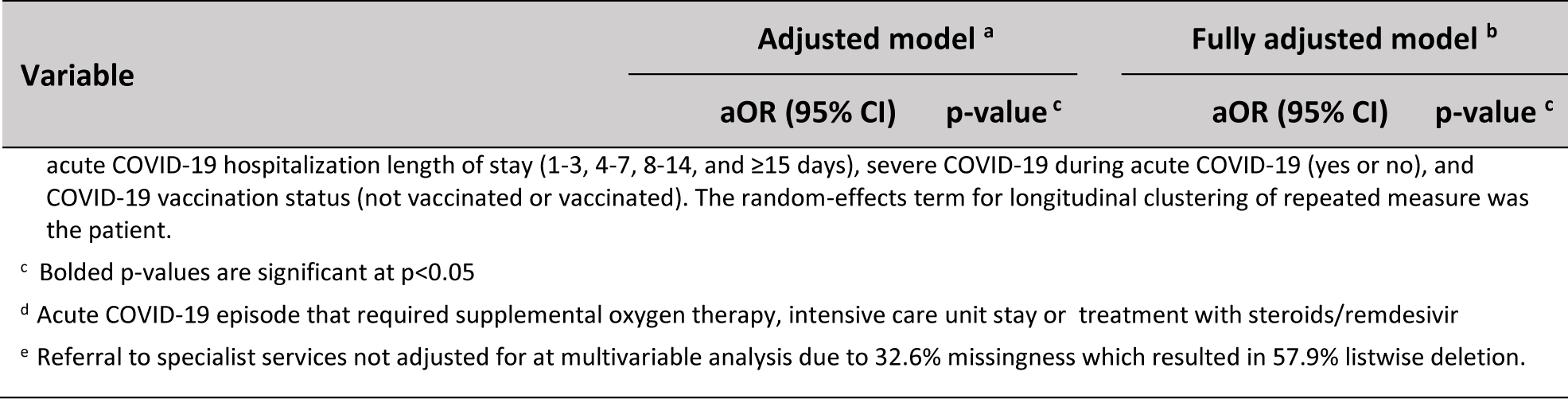
Factors longitudinally association with long COVID among acute COVID-19 hospitalized patients in Zambia, August 2020 – January 2023 (N=540)

In the longitudinal sub-analysis that included both those that were inpatients and outpatients during acute COVID-19 (S4 Table), similar associations were observed as to the cross section sub-analysis (S2 Table). The likelihood of presenting with long COVID significantly (p<0.001) declined, overall, across subsequent PAC-19 clinic visits (S4 Table). The presence of comorbidities were longitudinally associated with increased odds of long COVID by a factor of 1.67 (95% CI: 1.02-2.75). Hospitalized patients during acute COVID-19, vaccinated and male patients were associated with non-significant reduced likelihood of long COVID. Similarly, order age groups and presence of newly diagnosed medical conditions were associated with non-significant, however increased, odds of long COVID.

## Discussion

Long COVID symptoms were common among patients presenting for care in specialized post-acute COVID-19 clinics in Zambia, which could represent potential substantial morbidity from COVID-19. Factors associated with long COVID in Zambia included severe COVID-19, hospital length of stay and presence of pre-existing comorbidities. Hospitalization was not significantly associated with long COVID while patients with limitations in daily activities that may have impacted on the patients’ functional status were independently associated with long COVID. Longitudinally vaccination patients had indistinguishable likelihood of long COVID and so was sex and age.

A severe form of COVID-19 is known to increase the risk of an intense immune response that could lead to widespread inflammation [50]. The inflammatory response can persist post-acute infection and could have led to the immune system being dysregulated, leading to prolonged persistent symptoms and complications of COVID-19 in the patients. This could have led to observed association of severe COVID-19 long COVID as has been reported in other studies [35, 36, 51, 52].

Patients with longer hospital stays may have received intensive medical interventions and additional treatments. This could have potentially affected the immune response and the body’s ability to recover from the acute COVID-19p, which in turn may have increased the likelihood of long COVID. Furthermore, patients with pre-existing comorbidities may have been more susceptible to severe COVID-19 and may have been at higher risk of developing long COVID. This is consistent with what is reported by other studies on patients with longer hospitalization stay for acute COVID-19 or pre-existing comorbidities and their association to long COVID [52–54].

Patients referred to specialist services may have had residual organ damage from acute COVID-19 which may have required specialist care. This may have been closely related to the observed limitations in daily activities that may have impacted on the patients’ functional status since acute COVID-19 as have been reported in a previous study [9]. Although this association was independently significant, we could not adjust for it at multivariable analysis due to missingness that resulted in listwise deletion of more than half the study sample. Furthermore, no feedback systems were in place to inform PAC-19 clinics of the outcome of specialist care of referred patients.

COVID-19 vaccines have been shown to be effective at preventing symptomatic SARS-CoV-2 infections and long COVID [55, 56]. In this study, however, vaccinated patients failed to show significant protective association between COVID vaccination and long COVID. A possible explanation for this finding might be the heterogeneity in vaccination status during the study since Zambia only began offering COVID-19 vaccination in April 2021 (i.e., right before the delta wave and ∼8 months after patients began presenting for care in PAC-19 clinics) [57]. For example, among the patients diagnosed with SARS-CoV-2 during the wild type and beta dominant variants periods, prior to publicly available COVID-19 vaccination in Zambia, only 1 patient ever reported being vaccinated.

Unlike pre-existing comorbidities, newly diagnosed medical conditions had a non-significant association with long COVID. Newly diagnosed medical conditions in COVID-19 patients may have been as a result of multiorgan effects or autoimmune conditions [3]. Fewer patients, overall, had newly diagnosed medical conditions which could explain the non-significant association. Sex and age were similarly not associated with long COVID which seem to suggest that factors associated with long COVID in Zambia were largely clinical as opposed to demographic.

With regards to recovery time from long COVID, most patients with long COVID had symptom resolution by the second month of follow up. Overall, we found that commonly reported long COVID symptoms in Zambia were similar to findings in other studies [35, 36, 38, 52]. However, symptoms like forgetfulness (commonly reported among long COVID patients) and change in sleep were less commonly reported in our study compared to previous reports [17, 20].

In both the cross-sectional and longitudinal analysis, inpatients compared to outpatient care during COVID-19 episode were found to be associated with indistinguishable likelihood of long COVID. One reason for this might be that those who were outpatient and attending PAC-19 clinics were a special subset of outpatients with COVID-19 (i.e., they were seeking out or were referred for follow-up care during acute COVID-19e). Furthermore, inpatients (the majority of whom had severe COVID-19) might have been more likely to be referred to PAC-19 clinic whereas those who were outpatient were likely self-referral to PAC-19 clinic meaning only those who really needed it (i.e., those with ongoing symptoms) were in this study.

The findings in this study are subject to several limitations. Firstly, only patients attending PAC-19 clinics were included in the study, so other patients with long COVID who sought care elsewhere (or didn’t seek care) were not included. This potentially represents some form of selection bias within the study since other long COVID patients could have by-passed the PAC-19 clinic and presented directly to clinicians or specialist service at outpatient departments. Furthermore, PAC-19 clinics took time to scale-up throughout Zambia and some persons might still have faced difficulty accessing them. Overall, fewer than 1% of the over 344,000 confirmed COVID-19 cases in Zambia as of January 2023 presented for care in PAC-19 clinics.

Secondly, COVID-19 illness history (i.e., date of SARS-CoV-2 testing, hospitalization status, vaccination history, acute COVID-19 episode details, etc.) were self-reported by patients to clinicians which cannot rule out self-serving bias. Thirdly, the study utilized routinely collected clinical information from PAC-19 clinics which had substantial missingness for some key variables such as vaccination status and referral to specialist services. Thus, these variables were not adjusted for at multivariable analyses so as to maintain sample size. And lastly, patients were not retested for SAR-CoV-2 infection at subsequent PAC-19 review visits and so the possibility of reinfection could not be ruled out.

## Conclusion

Given the burden of COVID-19 in Zambia, the findings of a potentially substantial morbidity due to long COVID, and that yet few patients overall with COVID-19 had attended a PAC-19 clinic, scaling up PAC-19 care services and integrating into routine clinical care could improve access by patients and further aid in understanding the true burden of long COVID in Zambia. This could be coupled with efforts to expand knowledge of long COVD among clinicians and the general population in Zambia. Scaling of vaccination, may likely, provide better outcomes not only for long COVID but also during acute SARS-CoV-2 infection. Improving data quality for routine clinical data, could possibly be achieved by utilizing the existing electronic health record in Zambia (SmartCare) for all clinical encounters. Information sharing between referral care services and PAC-19 clinics could aid in understanding the outcome of long COVID among patients requiring specialist care.

### Disclaimer

*The findings and conclusions in this report are those of the authors and do not necessarily represent the official position of the US Centers for Disease Control and Prevention (CDC) or the study funders*.

## Data Availability

Data are not currently publicly available but may be obtained by a third party. The data are de-identified participant data, available with permission from the Government of the Republic of Zambia (GRZ) – Ministry of Health (MoH). A request to access the data can be made to the corresponding author, who will need to get permission from GRZ (MoH) to avail the data. Protocols and statistical analysis information is available as per above.

## Notes

### Competing Interest Statement

The authors have declared no competing interest.

### Funding Statement

This study has been supported in part by the President's Emergency Plan for AIDS Relief (PEPFAR) through the Centers for Disease Control and Prevention (CDC) Grant number/CoAg ID number GH002234. The study sponsor or funder had no role, in the study design collection, analysis, and interpretation of data writing of the report and the decision to submit the article for publication. In addition, there is independence of researchers from funders and all authors, external and internal, had full access to all of the data (including statistical reports and tables) in the study and can take responsibility for the integrity of the data and the accuracy of the data analysis.

### Author Declarations

The Ethics Committee of University of Zambia Biomedical Research gave waiver for informed consent (Ref No. 2711-2022) for this work. The Zambia National Health Research Authority approved this non-research work (Ref No: NHRA0002/26/05/2022).

### Summary of Updates

Included figures, tables and supplementary materials. Addressed reviewers' comments from a journal the paper is under review for publication.

