## Supplementary figures and images for "Clinical Characteristics and Factors Associated with Long COVID in Zambia, August 2020 to January 2023: A Mixed Methods Design"

### S1 Fig

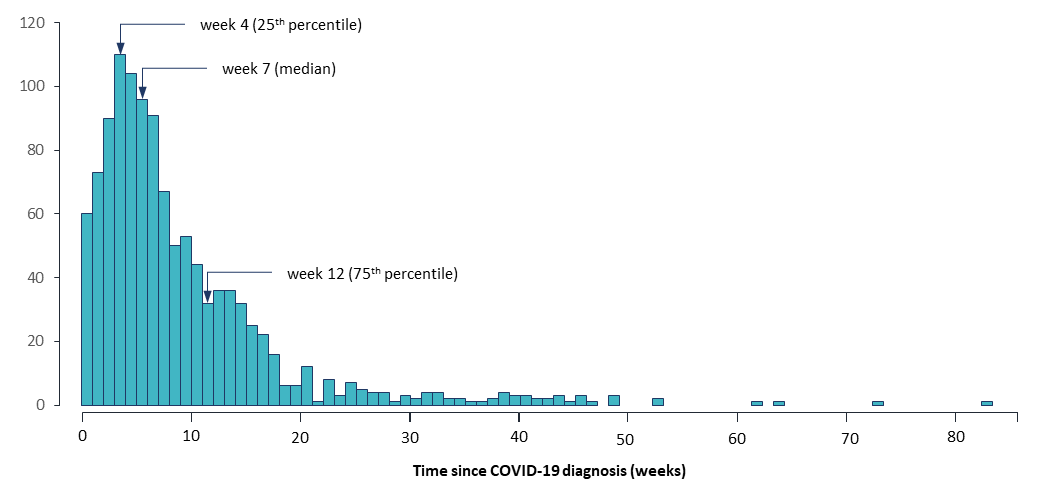
