## Supplementary material for "Clinical Characteristics and Factors Associated with Long COVID in Zambia, August 2020 to January 2023: A Mixed Methods Design": S1 Table

**S1 Table:** Frequencies of pre-existing and newly diagnosed comorbidities of PAC-19 Clinic Patients in Zambia, Aug. 2020‒Jan. 2023

| **Characteristic** | **Presence of long COVID symptoms** | | | **p-value ^a^** |
| --- | --- | --- | --- | --- |
|  | **Yes (222)**  **n (%)** | **No (432)**  **n (%)** | **Overall (654)**  **n (%)** |  |
| **Top five pre-existing comorbidities ^b^** |  |  |  |  |
| **Hypertension** |  |  |  | 0.095 |
| No | 55 (24.8) | 134 (31.0) | 189 (28.9) |  |
| Yes | 167 (75.2) | 298 (69.0) | 465 (71.1) |  |
| **Diabetes** |  |  |  | 0.525 |
| No | 167 (75.2) | 315 (72.9) | 482 (73.7) |  |
| Yes | 55 (24.8) | 117 (27.1) | 172 (26.3) |  |
| **HIV status** |  |  |  | 0.253 |
| Negative | 172 (77.5) | 317 (73.4) | 489 (74.8) |  |
| Positive | 50 (22.5) | 115 (26.6) | 165 (25.2) |  |
| **Cardiovascular disease** |  |  |  | 0.641 |
| No | 199 (89.6) | 382 (88.4) | 581 (88.8) |  |
| Yes | 23 (10.4) | 50 (11.6) | 73 (11.2) |  |
| **Obesity** |  |  |  | 0.421 |
| No | 212 (95.5) | 406 (94.0) | 618 (94.5) |  |
| Yes | 10 (4.5) | 26 (6.0) | 36 (5.5) |  |
| **Newly diagnosed medical conditions ^c^** |  |  |  |  |
| **Hypertension** |  |  |  | 0.913 |
| No | 20 (43.5) | 31 (42.5) | 51 (42.9) |  |
| Yes | 26 (56.5) | 42 (57.5) | 68 (57.1) |  |
| **Diabetes** |  |  |  | 0.780 |
| No | 22 (47.8) | 33 (45.2) | 55 (46.2) |  |
| Yes | 24 (52.2) | 40 (54.8) | 64 (53.8) |  |
| **Deep vein thrombosis/pulmonary embolism** | |  |  | >0.999 |
| Negative | 45 (97.8) | 72 (98.6) | 117 (98.3) |  |
| Positive | 1 (2.2) | 1 (1.4) | 2 (1.7) |  |
| **^a^** Bolded p-values are significant at p<0.05  ^b^ Denominator for pre-existing comorbidities are 654, i.e., patients with presence of comorbidities (see Table 1) ^c^ Denominator for newly diagnosed conditions is 119, i.e., patients with comorbidities detected at the time of SARS-CoV-2 diagnosis (see Table 1) | | | | |
