## Supplementary material for "Clinical Characteristics and Factors Associated with Long COVID in Zambia, August 2020 to January 2023: A Mixed Methods Design": S2 Table

**S2 Table:** Factors associated with long COVID at first visit to PAC-19 clinic among acute COVID-19 inpatients and outpatients in Zambia, August 2020 – January 2023 (N=1,359)

| **Variable** | **Unadjusted OR (95% CI)** | | | | **p-value ^a^** | **Adjusted OR (95% CI)^a^** | | **p-value ^a^** |
| --- | --- | --- | --- | --- | --- | --- | --- | --- |
| **Sex** | |  | | |  |  |  | |
| Female | | *Referent* | | |  | Referent | - | |
| Male | | 1.00 (0.79, 1.27) | | | 0.994 | 0.97 (0.74, 1.27) | 0.839 | |
| **Age group (years)** | |  | | |  |  |  | |
| ≤29 | | *Referent* | | |  | *Referent* | - | |
| 30-39 | | 1.59 (0.91, 2.84) | | | 0.113 | 1.30 (0.71, 2.44) | 0.400 | |
| 40-49 | | 1.99 (1.19, 3.44) | | | **0.011** | 1.79 (1.03, 3.22) | **0.044** | |
| 50-59 | | 1.67 (1.00, 2.88) | | | 0.057 | 1.30 (0.74, 2.36) | 0.367 | |
| 60+ | | 2.11 (1.30, 3.56) | | | **0.004** | 1.47 (0.86, 2.61) | 0.170 | |
| **Presence of comorbidities ^b^** | |  | | |  |  |  | |
| No | | *Referent* | | |  | *Referent* | - | |
| Yes | | 1.82 (1.43, 2.32) | | | **<0.001** | 1.55 (1.16, 2.08) | **0.003** | |
| **Presence of newly diagnosed medical conditions ^c^** | | |  | |  |  |  | |
| No | | *Referent* | | |  | *Referent* | - | |
| Yes | | 1.73 (1.17,2.55) | | | **0.006** | 1.35 (0.87, 2.06) | 0.173 | |
| **Hospitalization status during acute COVID-19** | | | |  |  |  |  | |
| Outpatient | | *Referent* | | |  | *Referent* | - | |
| Inpatient | | 1.16 (0.81, 1.69) | | | 0.428 | 1.05 (0.72, 1.55) | 0.794 | |
| **Vaccination status** | |  | | |  |  |  | |
| Not vaccinated | | *Referent* | | |  | *-* | - | |
| Vaccinated | | 0.90 (0.66, 1.23) | | | 0.520 | - | - | |
| **Referral to specialist Services ^d^** | |  | | |  |  |  | |
| No | | *Referent* | | |  | *-* | - | |
| Yes | | 2.34 (1.37, 4.00) | | | **0.002** | - | - | |
| Abbreviation: OR: odds ratio; CI: confidence interval  **^a^** Bolded p-values are significant at p<0.05  **^b^** Pre-existing comorbidity: hypertension, diabetes, cardiovascular disease, cancer, immunosuppression, chronic lung, kidney, and liver diseases, obesity, HIV and TB.  **^c^** Comorbidities (hypertension, diabetes, and HIV) diagnosed at the time of SARS CoV-2 infection.  **^d^** Vaccination status and referral to specialist services not adjusted for at multivariable analysis due to >10% missingness which resulted in 50.5% listwise deletion. | | | | | | | | |
