## Supplementary material for "Clinical Characteristics and Factors Associated with Long COVID in Zambia, August 2020 to January 2023: A Mixed Methods Design": S3 Table

**S3 Table:** Longitudinal Demographic and Clinical Characteristics of PAC-19 Clinic Patients in Zambia, Aug. 2020‒Jan. 2023

| **Characteristic** | **PAC-19 clinic visit** | | | | | **p-value ^a^** |
| --- | --- | --- | --- | --- | --- | --- |
|  | **Visit 1 (548)**  **n (%)** | **Visit 2 (548)**  **n (%)** | **Visit 3 (234)**  **n (%)** | **Visit 4 (115)**  **n (%)** | **Visit 5 (50)**  **n (%)** |  |
| **Prevalence of long COVID** |  |  |  |  |  | **<0.001** |
| No | 135 (24.6) | 255 (46.5) | 94 (40.2) | 58 (50.4) | 37 (74.0) |  |
| Yes | 413 (75.4) | 293 (53.5) | 140 (59.8) | 57 (49.6) | 13 (26.0) |  |
| **Sex** |  |  |  |  |  | 0.883 |
| Female | 287 (53.4) | 287 (53.4) | 120 (52.4) | 59 (51.3) | 30 (60.0) |  |
| Male | 250 (46.6) | 250 (46.6) | 109 (47.6) | 56 (48.7) | 20 (40.0) |  |
| (Missing) | 11 | 11 | 5 | 0 | 0 |  |
| **Age (years)** |  |  |  |  |  | 0.122 |
| Median (IQR) | 56 (46, 66) | 56 (46, 66) | 58 (48, 67) | 58 (49, 67) | 60 (50, 67) |  |
| (Missing) | 7 | 7 | 3 | 1 | 1 |  |
| **Age group (years)** |  |  |  |  |  | 0.804 ^b^ |
| <29 | 25 (4.6) | 25 (4.6) | 7 (3.0) | 2 (1.8) | 1 (2.0) |  |
| 30-39 | 53 (9.8) | 53 (9.8) | 20 (8.7) | 7 (6.1) | 2 (4.1) |  |
| 40-49 | 113 (20.9) | 113 (20.9) | 40 (17.3) | 21 (18.4) | 9 (18.4) |  |
| 50-59 | 134 (24.8) | 134 (24.8) | 58 (25.1) | 30 (26.3) | 11 (22.4) |  |
| 60+ | 216 (39.9) | 216 (39.9) | 106 (45.9) | 54 (47.4) | 26 (53.1) |  |
| (Missing) | 7 | 7 | 3 | 1 | 1 |  |
| **Dominant SARS-CoV-2 variant at diagnosis** ^c^ | | | |  |  | 0.152 ^b^ |
| Wild type | 34 (6.3) | 34 (6.3) | 21 (9.1) | 8 (7.0) | 4 (8.0) |  |
| Beta | 86 (15.9) | 86 (15.9) | 37 (15.9) | 10 (8.7) | 3 (6.0) |  |
| Delta | 331 (61.2) | 330 (61.0) | 147 (63.4) | 83 (72.2) | 37 (74.0) |  |
| Omicron | 90 (16.6) | 91 (16.8) | 27 (11.6) | 14 (12.2) | 6 (12.0) |  |
| (Missing) | 7 | 7 | 2 | 0 | 0 |  |
| **Time since diagnosis (weeks)** | | |  |  |  | **<0.001** |
| Median (IQR) | 4 (2, 6) | 7 (5, 11) | 10 (8, 14) | 13 (11, 18) | 16 (14, 22) |  |
| (Missing) | 112 | 114 | 42 | 15 | 6 |  |
| **Newly diagnosed medical conditions ^d^** | | |  |  |  | 0.953 |
| No | 488 (89.1) | 488 (89.1) | 207 (88.5) | 101 (87.8) | 46 (92.0) |  |
| Yes | 60 (10.9) | 60 (10.9) | 27 (11.5) | 14 (12.2) | 4 (8.0) |  |
| **Presence of comorbidities** ^e^ | | |  |  |  | **0.002** |
| No | 235 (42.9) | 235 (42.9) | 81 (34.6) | 33 (28.7) | 13 (26.0) |  |
| Yes | 313 (57.1) | 313 (57.1) | 153 (65.4) | 82 (71.3) | 37 (74.0) |  |
| **Hospitalization status during acute COVID-19** | | |  |  |  | 0.893 |
| Outpatient | 7 (1.3) | 7 (1.3) | 2 (0.9) | 1 (0.9) | 1 (2.0) |  |
| Inpatient | 540 (98.7) | 540 (98.7) | 232 (99.1) | 114 (99.1) | 49 (98.0) |  |
| (Missing) | 1 | 1 | 0 | 0 | 0 |  |
| **Hospitalization duration (weeks) ^f^** | |  |  |  |  | 0.137 |
| Median (IQR) | 9 (5, 18) | 9 (5, 18) | 11 (6, 21) | 10 (6, 21) | 10 (6, 19) |  |
| (Missing) | 180 | 180 | 71 | 25 | 11 |  |
| **Hospital length of stay (days) ^f^** | |  |  |  |  | 0.523 ^b^ |
| 1-3 | 49 (13.6) | 49 (13.6) | 13 (8.1) | 5 (5.6) | 2 (5.3) |  |
| 4-7 | 101 (28.1) | 101 (28.1) | 43 (26.7) | 25 (28.1) | 11 (28.9) |  |
| 8-14 | 94 (26.1) | 94 (26.1) | 44 (27.3) | 23 (25.8) | 11 (28.9) |  |
| ≥15 | 116 (32.2) | 116 (32.2) | 61 (37.9) | 36 (40.4) | 14 (36.8) |  |
| (Missing) | 180 | 180 | 71 | 25 | 11 |  |
| **Severe illness ^f^** |  |  |  |  |  | 0.918 |
| No | 189 (35.0) | 189 (35.0) | 79 (34.1) | 42 (36.8) | 17 (34.7) |  |
| Yes | 351 (65.0) | 351 (65.0) | 153 (65.9) | 72 (63.2) | 32 (65.3) |  |
| **Vaccination status** |  |  |  |  |  | **<0.001** |
| Not vaccinated | 421 (80.0) | 390 (74.1) | 161 (71.2) | 67 (58.3) | 21 (42.0) |  |
| Vaccinated | 105 (20.0) | 136 (25.9) | 65 (28.8) | 48 (41.7) | 29 (58.0) |  |
| (Missing) | 22 | 22 | 8 | 0 | 0 |  |
| **Referral to specialist services ^g^** | | |  |  |  | 0.161 |
| No | 299 (94.3) | 367 (92.2) | 152 (88.9) | 75 (90.4) | 35 (87.5) |  |
| Yes | 18 (5.7) | 31 (7.8) | 19 (11.1) | 8 (9.6) | 5 (12.5) |  |
| (Missing) | 231 | 150 | 63 | 32 | 10 |  |
| ^a^ Pearson Chi-square test was used for proportions and Wilcoxon rank sum test for medians unless otherwise stated. Bolded p-values are significant at p<0.05. Bonferroni correction for pairwise compariosns  ^b^ Fisher’s exact test  ^c^ SARS CoV-2 variants classification based on GISAID (2023) rather than sequenced specimens from study patients  ^d^ Pre-existing comorbidity: hypertension, diabetes, cardiovascular disease, cancer, immunosuppression, chronic lung, kidney, and liver diseases, obesity, HIV and TB.  **^e^** Comorbidities (hypertension, diabetes, and HIV) diagnosed at the time of SARS CoV-2 infection.  ^f^ Denominator is PAC-19 clinic visit specific patients hospitalized for acute COVID-19, i.e., visit 1: 540; visit 2: 540; visit 3: 232; visit 4: 114 and visit 5: 49.  ^g^ Specialist services included physiotherapy, cardiology, endocrinology, nephrology, psychiatry, and pulmonology | | | | | | |
