## Supplementary material for "Clinical Characteristics and Factors Associated with Long COVID in Zambia, August 2020 to January 2023: A Mixed Methods Design": S4 Table

**S4 Table:** Longitudinal risk factors and association for long COVID among acute COVID-19 hospitalized and non-hospitalized patients in Zambia, August 2020 – January 2023 (N=548)

| **Variable** |  | | **adjusted Model ^a^** | | **Fully adjusted model ^b^** | | |
| --- | --- | --- | --- | --- | --- | --- | --- |
|  |  | | **aOR (95% CI)** | **p-value ^c^** | **aOR (95% CI)** | | **p-value ^c^** |
| **PAC-19 clinic visit** | |  | |  |  |  | |
| 1 | | Referent | | - | Referent | - | |
| 2 | | 0.19 (0.13, 0.28) | | **<0.001** | 0.19 (0.13, 0.29) | **<0.001** | |
| 3 | | 0.09 (0.05, 0.18) | | **<0.001** | 0.06 (0.05, 0.22) | **<0.001** | |
| 4 | | 0.02 (0.01, 0.06) | | **<0.001** | 0.03 (0.01, 0.07) | **<0.001** | |
| 5 | | 0.00 (0.00, 0.01) | | **<0.001** | 0.00 (0.00, 0.01) | **<0.001** | |
| **Sex** | |  | |  |  |  | |
| Female | | *Referent* | | - | Referent | - | |
| Male | | 0.81 (0.52, 1.27) | | 0.367 | 0.84 (0.53, 1.34) | 0.460 | |
| **Age group (years)** | |  | |  |  |  | |
| ≤29 | | *Referent* | | - | Referent | - | |
| 30-39 | | 1.37 (0.67, 2.79) | | 0.208 | 2.13 (0.56, 8.08) | 0.266 | |
| 40-49 | | 1.05 (0.55, 2.01) | | 0.422 | 1.35 (0.40, 4.55) | 0.633 | |
| 50-59 | | 1.13 (0.59, 2.14) | | 0.438 | 1.30 (0.38, 4.37) | 0.676 | |
| 60+ | | 1.53 (0.82, 2.86) | | 0.094 | 1.99 (0.61, 6.56) | 0.256 | |
| **Presence of comorbidities** | |  | |  |  |  | |
| No | | *Referent* | | - | Referent | - | |
| Yes | | 1.86 (1.18, 2.92) | | **0.007** | 1.67 (1.02, 2.75) | **0.042** | |
| **Presence of newly diagnosed medical conditions** | |  | |  |  |  | |
| No | | *Referent* | | - | Referent | - | |
| Yes | | 1.55 (0.76, 3.14) | | 0.227 | 1.59 (0.76, 3.31) | 0.214 | |
| **Hospitalization status during acute COVID-19** | |  | |  |  |  | |
| Outpatient | | *Referent* | | - | Referent | - | |
| Inpatient | | 0.38 (0.05, 3.23) | | 0.379 | 0.36 (0.04, 2.97) | 0.342 | |
| **Vaccination status** | |  | |  |  |  | |
| Not vaccinated | | *Referent* | | - | Referent | - | |
| Vaccinated | | 0.80 (0.51, 1.25) | | 0.321 | 0.73 (0.46, 1.15) | 0.168 | |
| **Referral to specialist Services** ^d^ | |  | |  |  |  | |
| No | | *Referent* | | - | Referent | - | |
| Yes | | 0.99 (0.53, 1.84) | | 0.972 | - | - | |
| **Random Effects** | |  | |  |  |  | |
| Between PAC-19 clinic visit variance | |  | |  | 3.29 |  | |
| Within PAC-19 clinic visit variance | |  | |  | 3.97 |  | |
| Conditional intraclass correlation | |  | |  | 0.54 |  | |
| ^a^ Models adjusted for fixed effects term PAC-19 clinical visit occasion.  ^b^ Fully adjusted model included measurement occasion fixed effects term (PAC-19 clinic visit occasions) and patient level fixed effects terms: age (<29, 30-39, 40-49, 50-59, or +60 years), presence of comorbidities (yes or no), presence of newly diagnosed comorbidity (yes or no), acute COVID-19 hospitalization length of stay (1-3, 4-7, 8-14, and ≥15 days), severe illness during acute COVID-19 (yes or no), and COVID-19 vaccination status (not vaccinated or vaccinated). The random-effects term for longitudinal clustering of repeated measure was the patient.  ^c^ Bolded p-values are significant at p<0.05  ^d^ Referral to specialist services not adjusted for at multivariable analysis due to 32.5% missingness which resulted in 47.9% listwise deletion. | | | | | | | |
